# Epidemiological and Clinical Characteristics of 17 Hospitalized Patients with 2019 Novel Coronavirus Infections Outside Wuhan, China

**DOI:** 10.1101/2020.02.11.20022053

**Authors:** Jie Li, Shilin Li, Yurui Cai, Qin Liu, Xue Li, Zhaoping Zeng, Yanpeng Chu, Fangcheng Zhu, Fanxin Zeng

## Abstract

An increasing number of cases of novel coronavirus pneumonia (NCP) infected with 2019-nCoV have been identified in Wuhan and other cities in China, since December 2019. We analyzed data on the 17 confirmed cases in Dazhou to provide the epidemiologic characteristics of NCP outside Wuhan. Among them, 12 patients were still quarantined in the hospital, 5 patients were discharged NCP patients according to the national standards. Compared with non-discharged NCP patients, the discharged NCP patients had younger ages. Moreover, discharged NCP patients had higher heart rate, lymphocytes levels and monocytes levels than non-discharged NCP patients on admission to the hospital. Notably, all of 17 patients had abnormal increased C-reactive protein levels, and 16 patients had abnormal computed tomography images. This study provided some information that younger age, higher lymphocytes levels and monocytes levels at the diagnoses of 2019-nCoV may contributed to faster recovery and better therapeutic outcome.

## INTRODUCTION

An outbreak of acute respiratory illness, now named as novel coronavirus pneumonia (NCP)^1^, in Wuhan, Hubei, China since December 2019 has become a Public Health Emergency of International Concern (PHEIC)^2^. The pathogen of NCP is a 2019 novel coronavirus (2019-nCoV)^3^, possibly originated from wild animals in the Huanan Seafood Wholesale Market. Until 10 February 2020, a total number of 40, 261 NCP cases in China has been diagnosed, including 909 deaths^4^. Internationally, the patients of NCP have been confirmed in 25 countries and 5 continents^4^. Human-to-human transmission is the cause of most infections^5,6^. The main clinical manifestations of NCP patients are fever, cough, myalgia^5,7,8^. Notably, some cases have mild illness onset symptoms without fever. Acute respiratory distress syndrome (ARDS), RNAaemia, acute cardiac injury, and death could occur in severe patients^5^. However, there are few reports on the clinical manifestations of 2019-nCoV in cases or cured patients outside Wuhan. We aimed to report clinical characteristics of 17 cases with diagnosed 2019-nCoV infection in Dazhou, Sichuan.

## METHODS

Patients were admitted to the Dazhou Central Hospital from 22 January 2020 to 10 February 2020, with final follow-up for the study on 11 February 2020. Clinical data were collected from the hospital electronic record system. Suspected 2019-nCoV infection cases were hospitalized and quarantined. Sample of throat swab were obtained from the patient and sent to the Dazhou Center for Disease Control and Prevention to detect 2019-nCoV by applying quantitative polymerase chain reaction analysis^5^. Meanwhile, computed tomography was used for the suspected patients. The study was approved by the ethics committee of the Dazhou Central Hospital with written informed consent from the patients (IRB00000001-20001).

All statistical analyses were performed using SPSS version 20.0 (IBM, Armonk, NY, USA). The data between groups with variables were compared by χ2 test or Fisher’s exact test, and one-way analysis of variance followed by Tukey’s test. All means were reported with the corresponding standard deviation in tables. The *P* values < 0.05 were regarded as statistically significant.

## RESULTS

The median age of the 17 patients diagnosed with 2019-nCoV infection was 45 years (range from 22 to 65; 9 males and 8 females). Among them, 12 patients were still quarantined in the hospital (mean age, 49.4 years; range from 32 to 65; 5 males and 7 females), the remaining 5 patients were discharged NCP patients according to the national standards (mean age, 34.8 years; range from 22 to 48; 4 males and 1 females) (Table 1). Until 11 February 2020, no difference was observed in duration of hospitalization between non-discharged NCP patients and discharged NCP patients (*P*=0.852). Nine patients worked or visited Wuhan before the onset of the 2019-nCoV epidemic. Four patients had closely contacted with family members who diagnosed with 2019-nCoV infection. Two patients took the train that passed through Wuhan. Two patients had unknown epidemic area contact history (Table 1).

**Table 1.** Baseline Characteristics of patients with 2019-nCoV (N = 17) †.

|  | Overall (n=17) | Discharged<br>NCP patients<br>(n=5) | Non-discharged<br>NCP patients<br>(n=12) | *P* |
| --- | --- | --- | --- | --- |
| Age, y | 45.1 ± 12.8 | 34.8 ± 9.2 | 49.4 ± 11.7 | 0.026 |
| Male | 9 (52.9) | 4 (80.0) | 5 (41.7) | 0.169 |
| BMI | 25 ± 4.7 | 24.1 ± 3.5 | 25.3 ± 5.2 | 0.653 |
| **Cause of infection** |  |  |  |  |
| Live in Wuhan | 9 (52.9) | 4 (80.0) | 5 (41.7) | 0.294 |
| Via Wuhan | 2 (11.8) | 1 (20.0) | 1 (8.3) | 0.515 |
| None contact Wuhan | 2 (11.8) | 0 (0) | 2 (16.7) | 1.0 |
| Closely contacts<br>patients | 4 (23.5) | 0 (0) | 4 (33.3) | 0.261 |
| **Signs and symptoms** |  |  |  |  |
| Fever | 12 (70.6) | 4 (80.0) | 8 (66.7) | 1.0 |
| Febrile days, d | 6.2±3.5 | 5.3±3.3 | 6.6±3.8 | 0.554 |
| Fatigue | 8 (47.1) | 3 (60.0) | 5 (41.7) | 0.620 |
| Cough | 13 (76.5) | 3 (60.0) | 10 (83.3) | 0.538 |
| Dry cough | 7 (41.2) | 2 (40.0) | 5 (41.7) | 1.0 |
| Productive cough | 3 (17.6) | 0 (0) | 3 (25.0) | 0.516 |
| Rhinorrhea | 2 (11.8) | 0 (0) | 2 (16.7) | 1.0 |
| Myalgia | 4 (23.5) | 2 (60.0) | 2 (16.7) | 0.538 |
| Diarrhea | 2 (11.8) | 0 (0) | 2 (16.7) | 1.0 |
| Dizziness | 2 (11.8) | 2 (40.0) | 0 (0) | 0.074 |
| **Comorbidities** | 3 (17.6) | 2 (40.0) | 1 (8.3) | 0.191 |
| Hypertension | 1 (5.9) | 0 (0) | 1 (8.3) | 0.294 |
| Alcoholic hepatitis | 1 (5.9) | 0 (0) | 1 (8.3) | 0.294 |

*continue*
|  | Overall (n=17) | Discharged<br>NCP patients<br>(n=5) | Non-discharged<br>NCP patients<br>(n=12) | *P* |
| --- | --- | --- | --- | --- |
| Chronic pharyngitis | 1 (5.9) | 1 (20.0) | 0 (0) | 1.0 |
| Sinusitis | 1 (5.9) | 1 (20.0) | 0 (0) | 1.0 |
| Drinking history | 3 (17.6) | 1 (20.0) | 2 (16.7) | 0.276 |
| Smoking history | 3 (17.6) | 2 (40.0) | 1 (8.3) | 0.879 |
| Heart rate, bpm | 91.7±9.3 | 98.6±8.8 | 88.8±8.2 | 0.043 |
| Respiratory rate | 21.8±4.5 | 19.6±0.9 | 22.7±5.1 | 0.210 |
| hospital stay, d | 8.4±2.6 | 8.6±2.5 | 8.3±2.7 | 0.852 |

Eight of the twelve non-discharged NCP patients had a fever before hospitalized (mean±SD, 6.6±3.8 days). Other symptoms included cough (83.3%), fatigue (41.7%), myalgia (16.7%) and diarrhea (16.7%) (Table 1). Four of five discharged NCP patients had a fever before hospitalized (mean±SD, 5.3±3.3 days). Other symptoms including cough (60%), fatigue (60%), myalgia (60%) and dizziness (40%) (Table 1). Compared with non-discharged NCP patients, discharged NCP patients had higher heart rate (*P*=0.043), lymphocytes levels (*P*=0.005) and monocytes levels (*P*=0.019) on admission to the hospital (Table 1-2). All patients had abnormal increased C-reactive protein levels (Table 2).

**Table 2.**
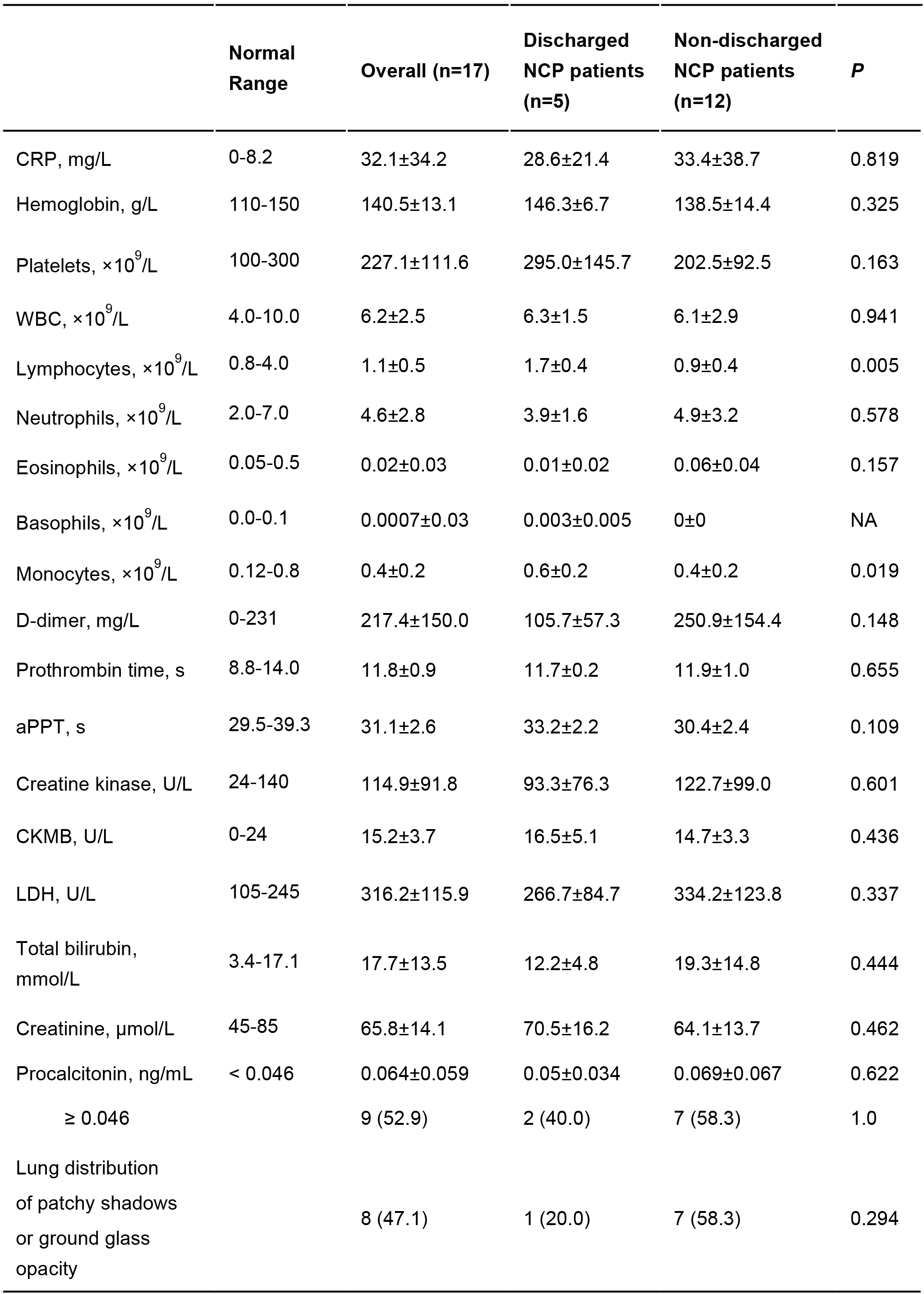
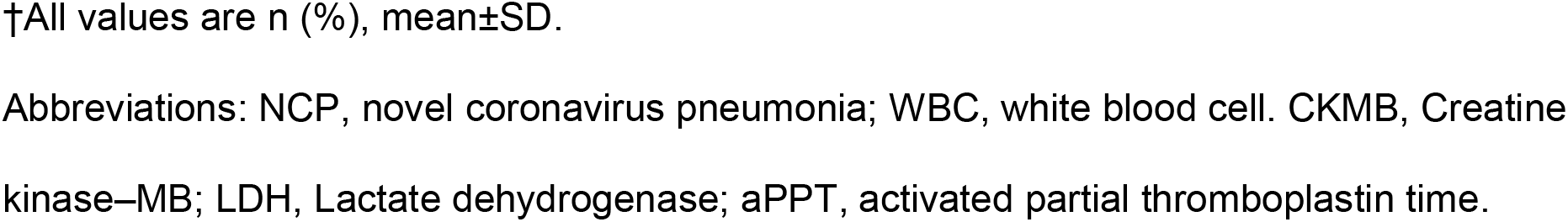
Laboratory Findings of Patients Infected With 2019-nCoV on Admission to Hospital (N = 17) †.

All seventeen patients had chest computed tomography at the admission of hospital. Seven non-discharged NCP patients showed patchy shadows or ground glass opacity (58.3%). Other five patients had either multiple interstitial changes in bilateral lobes or multilobed infectious lesions of both lungs (41.7%). Among discharged NCP patients, one patient had ground glass opacity (20%), three patients showed lung infections or inflammatory lesions (60%) and the youngest patients (22 years) had no abnormalities in the lungs (20%) (Figure).

**Figure.**
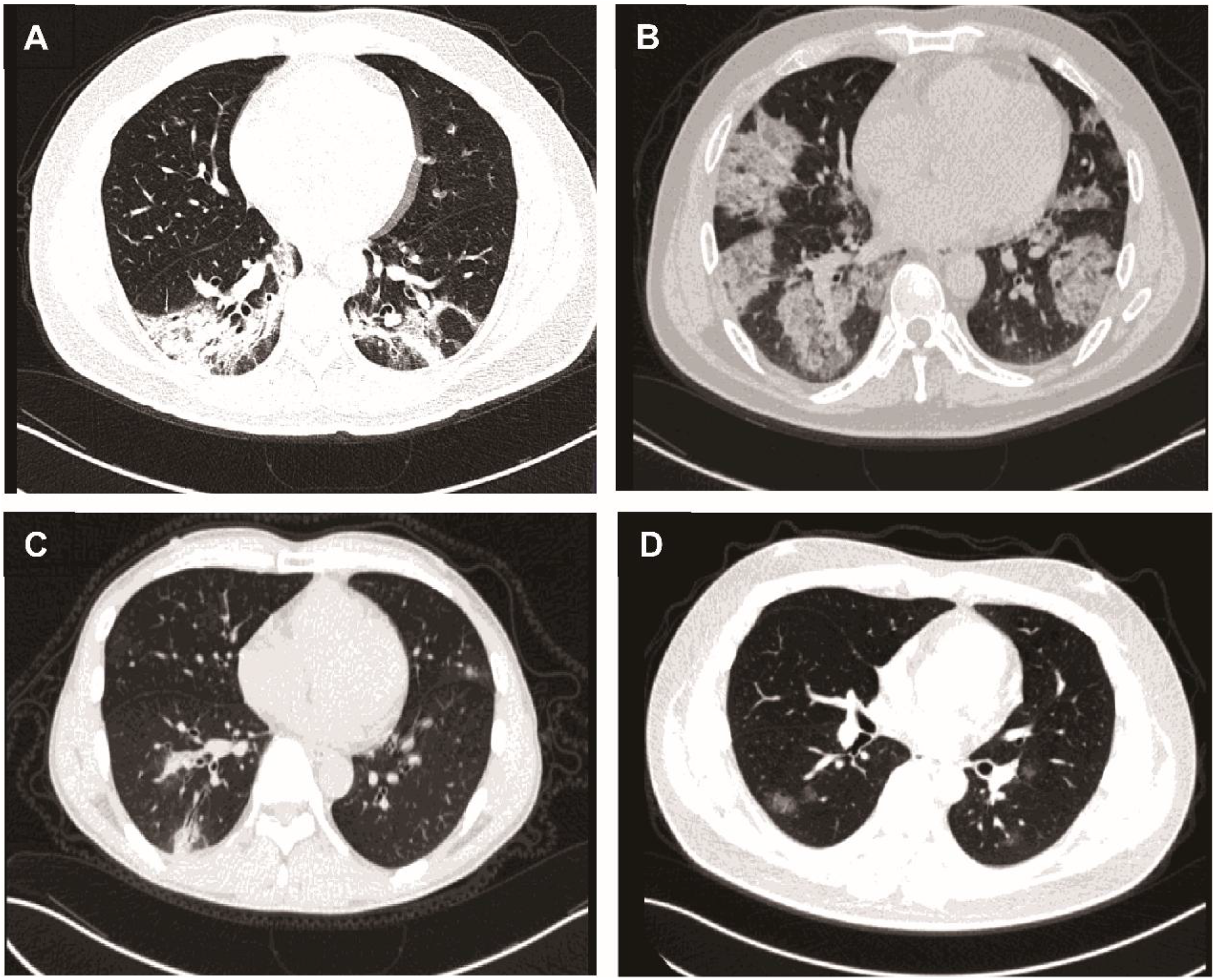
Chest Computed tomography (CT) images of patients infected with 2019-nCoV on admission to hospital. A, Chest CT scan obtained on February 2, 2020, from a 39-year-old man, showing bilateral ground glass opacities. B, Chest CT scan obtained on February 6, 2020, from a 45-year-old man, showing bilateral ground glass opacities. C, Chest CT scan taken on January 27, 2020, from a 48-year-old man (discharged after treatment on day 9), showing patchy shadows. D, Chest CT scan taken on January 23, 2020, from a 34-year-old man (discharged after treatment on day 11), showing patchy shadows.

## DISCUSSION

2019-nCoV is a betacoronavirus, like SARS and MERS, and it had infected hundreds of thousands of people since December 2019. There are currently no specific treatments available for 2019-nCoV. Therefore, the main treatment is optimized supportive care to reduce symptoms and maintain organ function in severe diseases^9,10^.

In our single-center case series of 17 hospitalized patients, it provided information on the epidemiology of the disease outside Wuhan. Routes of infection had no significant difference between discharged NCP patients and non-discharged NCP patients, but most patients had contact history with the epidemic area or close contacts with diagnosed patients. Two non-discharged NCP patients have been living long-term in Dazhou and have no contact history with epidemic area, which implied other possible infective routines.

In our study, the discharged NCP patients had younger ages compared with non-discharged NCP patients and no difference in gender distribution was observed. Our study was different from other reports which reveal that male are more likely to be infected^7^. Our data also suggested that age may be a risk factor for poor recovery. In addition, discharged NCP patients showed higher lymphocytes levels and monocytes levels on admission to hospital, which suggested immune deficiency nay be also a risk factor for poor outcome in patients. The result was inline with other study that higher level of lymphocyte was presented in the intensive care unit patients compared with that in non-intensive care unit patients^8^. Although C-reactive protein levels was abnormal increased in both discharged NCP and non-discharged NCP patients, no significant difference was presented in the two groups. Similar results was observed in lung distribution of patchy shadows or ground glass opacity. This study provided some information that younger age, higher lymphocytes levels and monocytes levels at the diagnoses of 2019-nCoV may contributed to faster recovery and better therapeutic outcome.

## Data Availability

Available one year after publication

## Author contributions

Fanxin Zeng and Fangcheng Zhu designed the study. Yurui Cai and Qin Liu contributed to the data collection. Shilin Li contributed to the data analysis and plotting. Jie Li and Zhaoping Zeng contributed to the drafted the manuscript. Xue Li and Yanpeng Chu contributed to the manuscript revision and submission. All authors read and approved the final manuscript.

## Acknowledgements

This study was funded by the National Natural Science Foundation of China (81902861), the Scientific Research Fund of Sichuan health and Health Committee (No. 18PJ361) and the Scientific Research Fund of Technology Bureau in Sichuan Province (No. 2018138, No. 2018JY0324).

We thank Fanwei Zeng, Qiliang Tan, Pingfei Wang,Lin Xu (Dazhou Central Hospital) and Xiuqin Zhang (Peking University) for supporting this work.

